# Safety and Immunogenicity of a 100 μg mRNA-1273 Vaccine Booster for Severe Acute Respiratory Syndrome Coronavirus-2 (SARS-CoV-2)

**DOI:** 10.1101/2022.03.04.22271830

**Authors:** Spyros Chalkias, Howard Schwartz, Biliana Nestorova, Jing Feng, Ying Chang, Honghong Zhou, Frank J. Dutko, Darin K. Edwards, David Montefiori, Rolando Pajon, Brett Leav, Jacqueline M. Miller, Rituparna Das

## Abstract

**Importance:** Due to the emergence of highly transmissible SARS-CoV-2 variants, evaluation of boosters is needed.

**Objectives:** Evaluate safety and immunogenicity of 100-µg of mRNA-1273 booster dose in adults.

**Design:** Open-label, Phase 2/3 study.

**Setting:** Multicenter study at 8 sites in the U.S.

**Participants:** The mRNA-1273 100-µg booster was administered to adults who previously received a two dose primary series of 100-µg mRNA-1273 in the phase 3 Coronavirus Efficacy (COVE) trial, at least 6 months earlier.

**Intervention:** Lipid nanoparticle containing 100-µg of mRNA encoding the spike glycoprotein of SARS-CoV-2 (Wuhan-HU-1).

**Main Outcomes and Measures:** Solicited local and systemic adverse reactions, and unsolicited adverse events were collected after vaccination. Primary immunogenicity objectives were to demonstrate non-inferiority of the neutralizing antibody (nAb) response against SARS-CoV-2 based on the geometric mean titer (GMTs) and the seroresponse rates (SRRs) (booster dose vs. primary series in a historical control group). nAbs against SARS-CoV-2 variants were also evaluated.

**Results:** The 100-µg booster dose had a greater incidence of local and systemic adverse reactions compared to the second dose of mRNA-1273 as well as the 50-µg mRNA-1273 booster in separate studies. The geometric mean titers (GMTs; 95% CI) of SARS-CoV-2 nAbs against the ancestral SARS-CoV-2 at 28 days after the 100-µg booster dose were 4039.5 (3592.7,4541.8) and 1132.0 (1046.7,1224.2) at 28 days after the second dose in the historical control group [GMT ratio=3.6 (3.1,4.2)]. SRRs (95% CI) were 100% (98.6,100) at 28 days after the booster and 98.1% (96.7,99.1) 28 days after the second dose in the historical control group [percentage difference=1.9% (0.4,3.3)]. The GMT ratio (GMR) and SRR difference for the booster as compared to the primary series met the pre-specified non-inferiority criteria. Delta-specific nAbs also increased (GMT fold-rise=233.3) after the 100-µg booster of mRNA-1273.

**Conclusions and Relevance:** The 100-µg mRNA-1273 booster induced a robust neutralizing antibody response against SARS-CoV-2, and reactogenicity was higher with the 100-µg booster dose compared to the authorized booster dose level in adults (50-µg). mRNA-1273 100-µg booster dose can be considered when eliciting an antibody response might be challenging such as in moderately or severely immunocompromised hosts.

Trial Registration: NCT04927065

**Key Points:** Question: What is the safety and immunogenicity of a booster dose of 100 µg of mRNA-1273 in adults who previously received the primary series of mRNA-1273?

Findings: In this open-label, Phase 2/3 study, the 100 µg booster dose of mRNA-1273 had a greater incidence of local and systemic adverse reactions compared to a 50 µg booster dose of mRNA- 1273 or after the second dose of mRNA-1273 during the primary series. The 100 µg booster dose of mRNA-1273 induced a robust antibody response against the ancestral SARS-CoV-2 and variants.

Meaning: mRNA-1273 100 µg booster dose might be considered when eliciting an antibody response might be challenging, such as in moderately or severely immunocompromised hosts.

## INTRODUCTION

Since late 2019, the Severe Acute Respiratory Syndrome Coronavirus-2 (SARS-CoV-2) has infected hundreds of millions of people with millions of fatalities^1^ globally. mRNA-1273, the vaccine encoding the SARS-CoV-2 spike protein of the Wuhan-HU-1 isolate, has shown to be efficacious and well tolerated. In the phase 3 Coronavirus Efficacy (COVE) trial, the vaccine efficacy was 93.2% after a median follow-up of 5.3 months following a 2-dose primary series^2^.

The mRNA-1273 vaccine has been administered to hundreds of millions of adults globally. Following the initial emergency use authorization of the two-dose series, moderately or severely immunocompromised individuals were advised to also receive a third 100 µg dose, one month after the second dose based on enhanced immunogenicity with a 3-dose immunization in the immunocompromised population^3, 4^.

Multiple infection waves have occurred throughout the SARS-CoV-2 pandemic as a result of emerging SARS-CoV-2 variants that exhibit enhanced transmissibility and have the potential to circumvent immunity elicited by natural infection or vaccination over time^5, 6^. In 2021, the Delta variant (B.1.617.2) became dominant in multiple regions and caused breakthrough infections and disease, especially in individuals who were not recently immunized^5^. Vaccine authorization was expanded in many countries to include a 50 µg mRNA- 1273 booster dose, at least six months after completion of the primary immunization, to boost the immune response against SARS-CoV-2^7^. A booster dose has been shown to further prevent COVID-19 and the durability of the booster dose effectiveness is being evaluated.^6, 8, 9^

The more recently emergent Omicron variant (B.1.529) was first detected in November 2021 and is substantially divergent given the presence of >30 mutations in the spike protein, 15 of which are clustered in the receptor binding domain, the main target of neutralizing antibodies. The Omicron variant is highly transmissible, has the potential to escape immunity and is currently causing a wave of infections associated with morbidity and mortality worldwide^10, 11^. Initial assessment of the Omicron-specific neutralizing antibodies in sera from recipients of the 50 µg mRNA-1273 booster dose, using a pseudovirus assay, showed higher neutralizing antibody titers after the booster dose, compared to the Omicron-specific antibody response after the mRNA-1273 primary series^12^. Nonetheless, emerging vaccine effectiveness data show decreased effectiveness of a booster dose against infection from the Omicron variant compared to that against the Delta variant^6^.

As sequential waves of SARS-CoV-2 variants pose an ongoing public health risk, we initiated a phase 2/3 study to evaluate the safety, immunogenicity, and durability of different booster vaccines, including a higher booster dose (100 µg) of the prototype mRNA-1273 vaccine and variant-matched booster vaccines. Here we summarize interim analysis results on the safety, reactogenicity and immunogenicity one month after the booster dose of the 100 µg mRNA-1273.

## METHODS

### Study design

This is Part B of an ongoing open-label, Phase 2/3 study (mRNA-1273-P205; NCT04927065) to evaluate the safety, reactogenicity and immunogenicity of the 100 µg mRNA- 1273 vaccine when administered as a single booster dose to adult participants who received a primary series of 2 doses of 100 µg mRNA-1273 in the phase 3 COVE study^2, 13^, at least 6 months earlier (Figure 1).

**Figure 1:**
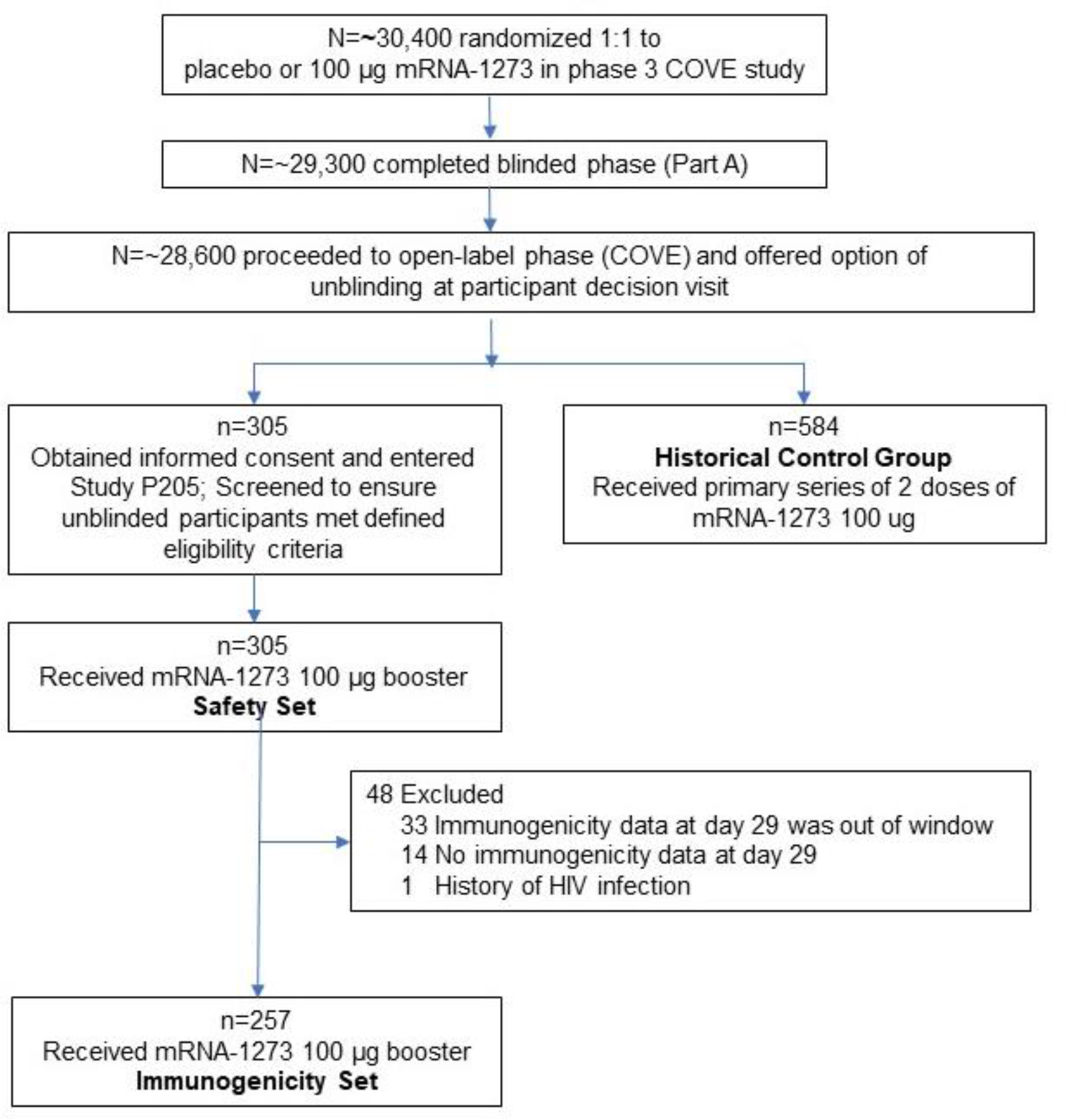
CONSORT Flow Diagram Trial profile. Participants who received two doses of mRNA-1273 in Phase 3 COVE trial were offered a booster injection of 100 μg of mRNA-1273 in this Phase 2/3, Part B, study. Eligibility to receive the booster included participants who completed 6 months of follow-up after the last injection received in the Phase 3 COVE trial. The screen failure rate will be presented for the entire study at the end-of-study analysis.

### Trial participants

Approximately 300 participants age 18 and older, who were actively enrolled and compliant in the phase 3 COVE trial, and who had completed their primary series of mRNA- 1273 at least 6 months prior, were enrolled from August 3-16, 2021, across 8 clinical sites in the United States. Upon meeting eligibility criteria and providing consent to enroll into the current Phase 2/3 trial, participants received a single booster dose of 100 µg mRNA-1273 on Day 1.

Participants who had known history of SARS-CoV-2 infection in the COVE trial or have had significant exposure to someone with SARS-CoV-2 infection or COVID-19 in the 14 days prior to screening, were excluded.

### Trial vaccine

The mRNA-1273 booster dose was a lipid nanoparticle containing mRNA encoding the spike glycoprotein of the Wuhan-HU-1 isolate of SARS-CoV-2.^7^ The booster dose of mRNA-1273 was administered at a dose level of 100 µg mRNA in a 0.5 mL volume.

### Study outcomes

The primary safety objective was to evaluate the safety and reactogenicity of a 100 µg mRNA-1273 administered as a single booster dose. The primary safety endpoints were solicited local and systemic adverse reactions (ARs) as recorded daily by participants during the 7 days after administration of the booster injection. Unsolicited treatment-emergent adverse events (TEAEs) were recorded by the study sites for a period of 28 days post-booster administration. Serious adverse events (SAEs), medically-attended adverse events (MAAEs) and AEs of special interest (AESIs) are being recorded by the study sites for the entire study period (approximately 12 months).

The primary immunogenicity objective was to demonstrate a non-inferior immune response against the ancestral SARS-CoV-2 with the D614G mutation at 28 days after the single booster dose of 100 µg mRNA-1273 compared with the immune response 28 days after the second of 2 priming doses of 100 µg mRNA-1273 in a historical control group. The historical control group consisted of 584 participants who received a primary series of 2 doses of mRNA- 1273 100 µg and were included in the random sub-cohort for Immunogenicity of the phase 3 COVE trial^2, 13, 14^. Random selection was performed for the historical control group to have a similar age distribution with the entire COVE study population (75% participants were ≥ 18 and <65 years old, and 25% participants were ≥65 years old)^14^. The details of the immunogenicity assays and the statistical analysis are in the Supplementary Appendix. Data from an separate phase 2 study (mRNA-P201, NCT04405076) where participants received a 50 µg mRNA-1273 booster dose are included^7^.

## RESULTS

### Trial Population

Three hundred and five participants received the mRNA-1273 100 µg booster dose and were included in the Safety Set. Two study sites were impacted by a hurricane which led to the inability to obtain post-vaccination blood samples at Day 29 within the per-protocol time window and hence 257 participants were included in the Immunogenicity Set (Table 2). The baseline demographics and characteristics of the recipients of the 100 µg mRNA-1273 booster dose were comparable to the historical control group. The mean ages (range) of the booster dose recipients were 54.2 (21-84) years in the booster group, and 52.1 (18-87) years in the historical control group. The percentage of participants who were White in the booster group was 92% and 72% in the historical control group; the corresponding rates for Hispanic and Latino was 22% and 31% respectively. The median (range) of the duration between the second dose of mRNA-1273 in the primary series and the booster dose was 326 (266-354) days in the Safety Set and 327 (266-354) days in the Immunogenicity Set. Five percent (14 of 305) of participants who received the booster dose had evidence of a prior SARS-CoV-2 infection, detected either via anti-nucleocapsid antibody detection in blood or polymerase chain reaction (PCR) nucleic acid testing in nasopharyngeal swabs prior to the administration of the booster dose.

**Table 1.** Baseline demographics and characteristics of the safety set, immunogenicity set, and the historical control group participants from the phase 3 COVE trial. RT-PCR = reverse transcription polymerase chain reaction. ^a^Percentages based on the number of participants in the safety set or the immunogenicity set. ^b^ Elecsys assay for binding antibody to SARS-CoV-2 nucleocapsid. ^c^Baseline SARS-CoV-2 status was positive if there was immunologic or virologic evidence of prior Covid-19, defined as positive binding antibody against SARS-CoV-2 nucleocapsid or positive RT-PCR test above the lower limit of quantification at day 1 (pre-booster for mRNA-1273-P205); Negative was defined as negative binding antibody against SARS-CoV-2 and a negative RT-PCR test day 1.

|  | Safety Set | Immunogenicity Set | Historical control group,<br>100 µg mRNA-1273 |
| --- | --- | --- | --- |
| Characteristics n (%)<sup>a</sup> | Recipients of 100 µg mRNA-1273<br>Booster Dose<br>N=305 | Recipients of 100 µg mRNA-1273<br>Booster Dose<br>N=257 | N=584 |
| Age at Screening (yr) |  |  |  |
| Mean (range) | 54.2 (21-84) | 54.7 (21-84) | 52.1 (18-87) |
| Age subgroup |  |  |  |
| ≥18 and <65 years | 216 (71) | 180 (70) | 438 (75) |
| ≥65 years | 89 (29) | 77 (30) | 146 (25) |
| Sex |  |  |  |
| Male | 174 (57) | 145 (56) | 308 (53) |
| Female | 131 (43) | 112 (44) | 276 (47) |
| Ethnicity |  |  |  |
| Hispanic or Latino | 66 (22) | 58 (23) | 183 (31) |
| Not Hispanic or Latino | 237 (78) | 197 (77) | 398 (68) |
| Not reported or unknown | 2 (1) | 2 (1) | 3 (1) |
| Race |  |  |  |
| White | 279 (92) | 237 (92) | 419 (72) |
| Black or African American | 17 (6) | 14 (5) | 109 (19) |
| Asian | 6 (2) | 4 (2) | 13 (2) |
| American Indian or Alaska Native | 0 | 0 | 8 (1) |
| Native Hawaiian or Other Pacific Islander | 0 | 0 | 3 (1) |
| Multiracial | 1 (<1) | 1 (<1) | 11 (2) |
| Other | 0 | 0 | 16 (3) |
| Not reported or unknown | 2 (1) | 1 (<1) | 5 (1) |
| Body Mass Index, (kg/m<sup>2</sup>) |  |  |  |
| n | 305 | 257 | 581 |
| Mean (SD) | 29.8 (6.5) | 29.9 (6.5) | 31.1 (7.9) |
| Duration between second injection of mRNA-1273 in Phase 3 COVE trial and the booster (days) |  |  |  |
| n | 305 | 257 | NA |
| Median | 326 | 327 |  |
| Range | 266-354 | 266-354 |  |
| Pre-booster/Baseline RT-PCR SARS-CoV-2 Results |  |  |  |
| Negative | 300 (98) | 254 (99) | 584 (100) |
| Positive | 5 (2) | 3 (1) | 0 |
| Missing | 0 | 0 | 0 |
| Pre-booster/Baseline antibody to SARS-CoV-2 nucleocapsid <sup>b</sup> |  |  |  |
| Negative | 295 (97) | 247 (96) | 584 (100) |
| Positive | 10 (3) | 10 (4) | 0 |
| Pre-booster/Baseline SARS-CoV-2 status <sup>c</sup> |  |  |  |
| Negative | 291 (95) | 245 (95) | 584 (100) |
| Positive | 14 (5) | 12 (5) | 0 |

**Table 2.** Neutralizing antibody titers (pseudovirus assay; ID50; pre-vaccination baseline titers) after the 100 µg booster dose of mRNA-1273. GM=geometric mean, CI=confidence interval, NE=not estimated, GMFR=geometric mean fold rise, GLSM=geometric least squares mean, GMR=geometric mean ratio. ^a^Number of participants with non-missing data at the corresponding timepoint. ^b^The 95% CI was calculated based on the t-distribution of the log-transformed values or the difference in the log-transformed values for GM value and GM fold-rise, respectively, and then back transformed to the original scale for presentation. ^c^Seroresponse at a participant level was defined as a change from below the LLOQ to equal or above 4 x LLOQ if the baseline was below the LLOQ, or at least a 4-fold rise if the baseline was equal to or above the LLOQ. For mRNA1273-P205 subjects without pre-Dose 1 antibody titer information and have corresponding Day 29 post-boost assessment, seroresponse is defined as >= 4*LLOQ for subjects with negative SARS-CoV-2 status at their pre-dose 1 of primary series, and these subjects’ antibody titer are imputed as <LLOQ at pre-dose 1 of primary series. For subjects who are without SARS-CoV-2 status information at pre-dose 1 of primary series, their pre-booster SARS-CoV-2 status is used to impute their SARS-CoV-2 status at their pre-dose 1 of primary series. For mRNA1273-P301 subjects without pre-Dose 1 antibody titer information for variant B.1.1.617 and have corresponding Day 57 B.1.1.617 assessment, seroresponse is defined as >= 4*LLOQ for subjects with negative SARS-CoV-2 status at their pre-dose 1 of primary series, and these subjects’ antibody titer are imputed as <LLOQ at pre-dose 1 of primary series. ^d^The number of participants meeting the criterion at the time point. Percentages are based on N1. ^e^The 95% CI was calculated using the Clopper-Pearson method. ^f^ The 95% CI was calculated using the Miettinen-Nurinen (score) confidence limits. ^g^The log-transformed antibody levels were analyzed using an analysis of covariance (ANCOVA) model with the treatment variable as fixed effect, adjusting for age group (<65, >=65 years). The treatment variable corresponded to the historical control group (randomly selected group from the Phase 3 COVE trial of the mRNA-1273 100 µg primary series) and study arm. The resulted LS means, difference of LS means, and 95% CI were back transformed to the original scale for presentation. For calculation of GMTs and GMFRs, antibody values reported as below the lower limit of quantification (LLOQ) were replaced by 0.5 times the LLOQ. Values that were greater than the upper limit of quantification (ULOQ) were converted to the ULOQ if actual values were not available. Missing results were not imputed.

|  | Ancestral SARS-CoV-2 with D614G |  | Delta (B.1.617.2) |  |
| --- | --- | --- | --- | --- |
|  | Recipients of 100 µg mRNA-1273 Booster Dose<br>N=257 | Historical control group<br>100 µg mRNA-1273<br>N=584 | Recipients of 100 µg mRNA-1273 Booster Dose<br>N=257 | Historical control group<br>100 µg mRNA-1273<br>N=584 |
| Pre-vaccination Baseline n<sup>a</sup> | 256 | 584 | 255 | 580 |
| GM titer (95% CI)<sup>b</sup> | 9.3<br>(NE-NE) | 9.7<br>(9.3-10.1) | 9.3<br>(NE-NE) | 9.3<br>(NE, NE) |
| 28 days after booster or 2<sup>nd</sup> dose n<sup>*</sup> | 257 | 584 | 257 | 580 |
| GM titer (95% CI)<sup>b</sup> | 4039.5<br>(3592.7-4541.8) | 1132.0<br>(1046.7-1224.2) | 2164.4<br>(1915.0-2446.3) | 383.1<br>(352.4-416.4) |
| GMFR (95% CI)<sup>b</sup> | 440.2<br>(391.7-494.6) | 116.8<br>(107.2-127.3) | 233.3<br>(206.4, 263.7) | 41.4<br>(38.1, 45.0) |
| Seroresponse rate<sup>c</sup><br>n/N1 (%)<sup>d</sup><br>(95% CI)<sup>e</sup> | 256/256 (100)<br>98.6-100 | 573/584 (98.1)<br>(96.7-99.1) | 255/255 (100)<br>(98.6-100) | 558/580 (96.2)<br>(94.3-97.6) |
| Difference in seroresponse rate after booster compared to Phase 3 COVE [% (95% CI)<sup>f</sup>] | 1.9<br>(0.4, 3.3) |  | 3.8<br>(2.3, -5.7) |  |
| GLSM (28 days after booster / 28 days after 2<sup>nd</sup> dose in Phase 3 COVE trial (95% CI)<sup>g</sup> | 3856.6<br>(3418.1-4351.4) | 1068.4<br>(980.4-1164.2) | 2074.0<br>(1825.6-2356.2) | 363.4<br>(331.8-397.9) |
| GMR (95% CI) | 3.6<br>(3.1-4.2) |  | 5.7<br>(4.9-6.6) |  |

### Safety

The median follow-up time after administration of the 100 µg booster dose was 66 days.

The incidences of solicited local or systemic adverse reactions within seven days after the 100 µg booster injection, the second dose of the primary series for the historical control group and after a 50 μg dose in a separate phase 2 study (mRNA-P201, NCT04405076) are shown in Figure 2 and in Supplementary Table 1^2, 7^ . The most common solicited local adverse reaction following the 100 µg booster dose was injection-site pain which occurred in 92.7% of participants and was reported as mostly mild in severity (Grade 1, 66.6%). The most common solicited systemic reactions in recipients of the 100 µg booster dose were fatigue (72.8%), myalgia (67.5%), and headache (61.9%). The majority of solicited local and systemic reactions were grade 1 (31.4%) or grade 2 (47.5%); 16.8% of reactions were grade 3, and there were no grade 4 reactions.

**Figure 2:**
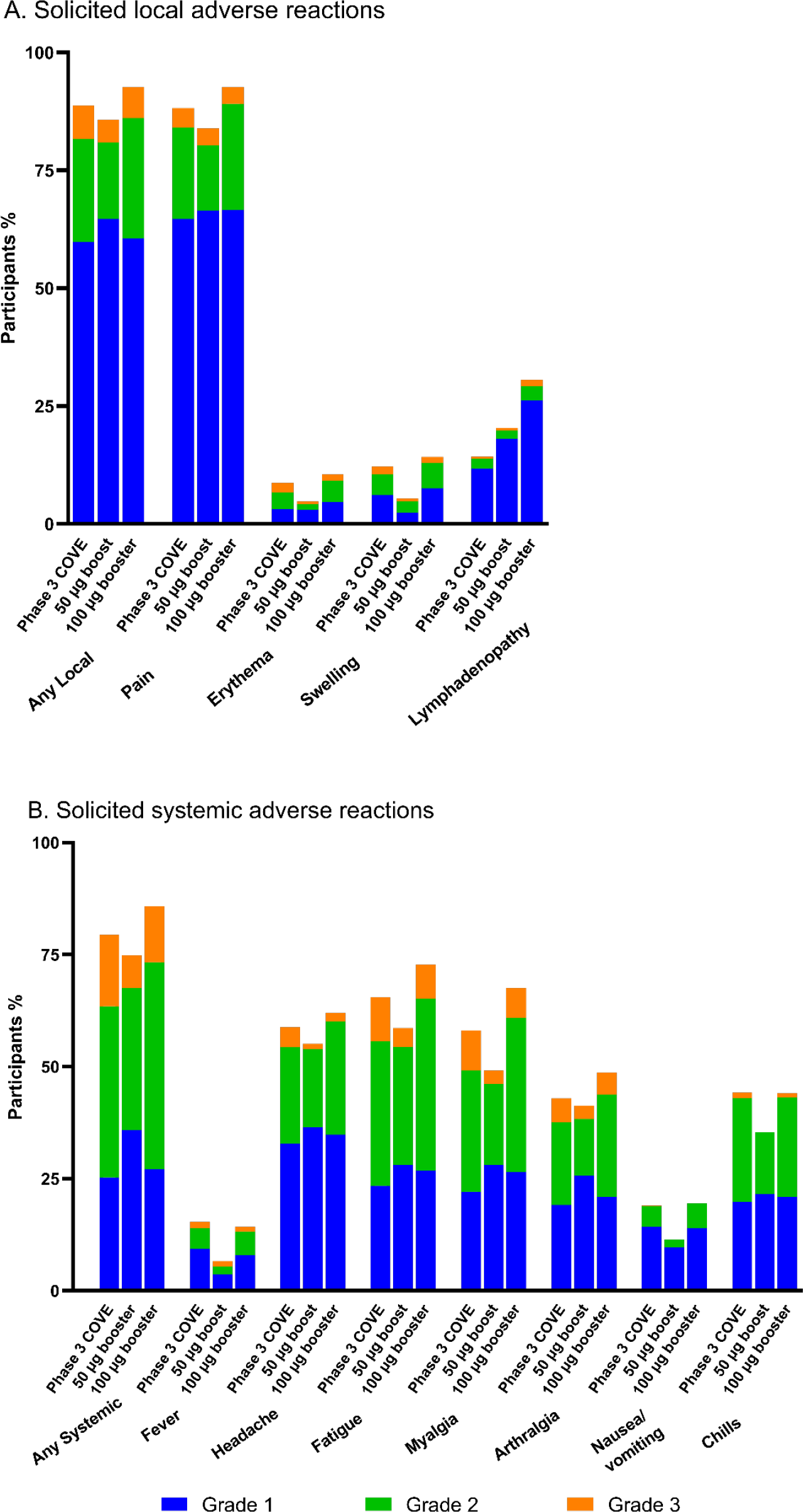
Solicited Local and Systemic Adverse Reactions. Shown are the percentages of participants who had a solicited local or systemic adverse reaction within 7 days after the second injection of 100 µg of mRNA-1273 of the primary series (n=14691; Phase 3 COVE trial), after the injection of a mRNA-1273 booster 50 µg dose (n=167; Phase 2 study mRNA-1273-P201) or after the injection of a 100 µg mRNA-1273 booster dose (n=303; the study reported here). In the phase 3 COVE trial, there were Grade 4, solicited systemic reports of fever (13/14,682; <0.1%) or nausea/vomiting (1/14,687; <0.1%) (not shown) whereas there were no Grade 4 adverse reactions with the booster doses.

Overall, the 100 µg booster dose had a numerically higher incidence of local and systemic adverse reactions compared to the second dose of the primary mRNA-1273 series and the 50 µg booster dose (Figure 2; Supplementary Table 1). There was a higher frequency of axillary swelling or tenderness after the 100 µg booster injection (30.5%) compared to that after the second dose of the 100 µg mRNA-1273 primary series in the phase 3 COVE trial (14.2%) and after the 50 µg booster injection (20.4%) in the separate phase 2 study (mRNA- P201). There also was a trend towards higher frequencies of solicited systemic reactions with the 100 µg booster dose compared to 50 µg booster dose, including the frequencies of fever (14.2% vs. 6.6%), fatigue (72.8% vs. 58.7%) and myalgia (67.5% vs. 49.1%). However, the incidence and severity of these reactions were comparable to what was observed after the primary series.

Unsolicited adverse events regardless of the relationship to the vaccination, and unsolicited events that were considered related to the vaccination by the investigators, up to 28 days after the 100 µg booster injection occurred in 16.4% (50 of 305) and in 5.9% (18 of 305) of participants, respectively. The most common unsolicited adverse events were fatigue (1.3%) and headache (1.0%). There were 6 serious adverse events, including one fatal adverse event of cardiopulmonary arrest nine days after the booster in a 72-year-old male with history of atherosclerosis. An autopsy was performed, and the cause of death was reported to be atherosclerotic and hypertensive cardiovascular disease. The event was deemed as unrelated to vaccination by the investigator. From the other 5 serious adverse events, one event of cerebrovascular accident (stroke), four days after the booster dose, was considered related to vaccination by the investigator. The event occurred in a 70-year-old male with history of coronary and peripheral vascular disease and subsequently clinically resolved. The events of cardiopulmonary arrest and cerebrovascular accident (stroke) were also reported as adverse events of special interest. Lastly, there were four asymptomatic (4/291, 1.4%) SARS-CoV-2 infections captured either via PCR (one participant) or anti-nucleocapsid antibody testing (three participants) at a routine testing time point after the first two weeks following the 100 µg booster injection among 291 participants who had history of SARS-CoV-2 infection prior to enrollment. There were no COVID-19 events (no symptomatic SARS-CoV-2 infections).

### Immunogenicity

Administration of a booster dose of 100 µg mRNA-1273 to participants who previously received the 2-dose primary series of 100 µg mRNA-1273, increased neutralizing antibodies to the ancestral (Wuhan-Hu-1) SARS-CoV-2 spike protein containing the D614G amino acid substitution (Table 2; Figure 3A; Supplementary Figure 1; Supplementary Tables 2-4). The geometric mean antibody titers (GMTs, 95% CIs) pre-vaccination (before Dose 1 of the primary series) were 9.3 (not estimated, [NE]) and increased to 4039.5 (3592.7,4541.8) at 28 days after the booster dose. The geometric mean fold rise (GMFR, 95% CIs) for the GMTs at 28 days after the booster compared to pre-vaccination was 440.2 (391.7-494.6) and GMFR (95% CI) compared to the pre-booster was 45.0 (38.9, 52.1). In comparison, the GMT (95% CIs) was 1132.0 (1046.7,1224.2) in the historical control group at 28 days after the second dose. The estimated geometric mean titer ratio (GMR; 95% CI) of the titers at 28 days after the booster dose compared to 28 days after the second 100 µg dose was 3.6 (3.1,4.2) and the GMR met the pre-specified non-inferiority margin of 1.5 (the lower bound of 95% CI was greater than 0.67). The GMTs 28 days after the 100 µg mRNA-1273 booster were also higher compared to the GMTs after the 50 µg mRNA-1273 booster [1951.7 (1729.6, 2202.4)] evaluated in the separate phase 2 study (mRNA-1273-P201).^15^

**Figure 3:**
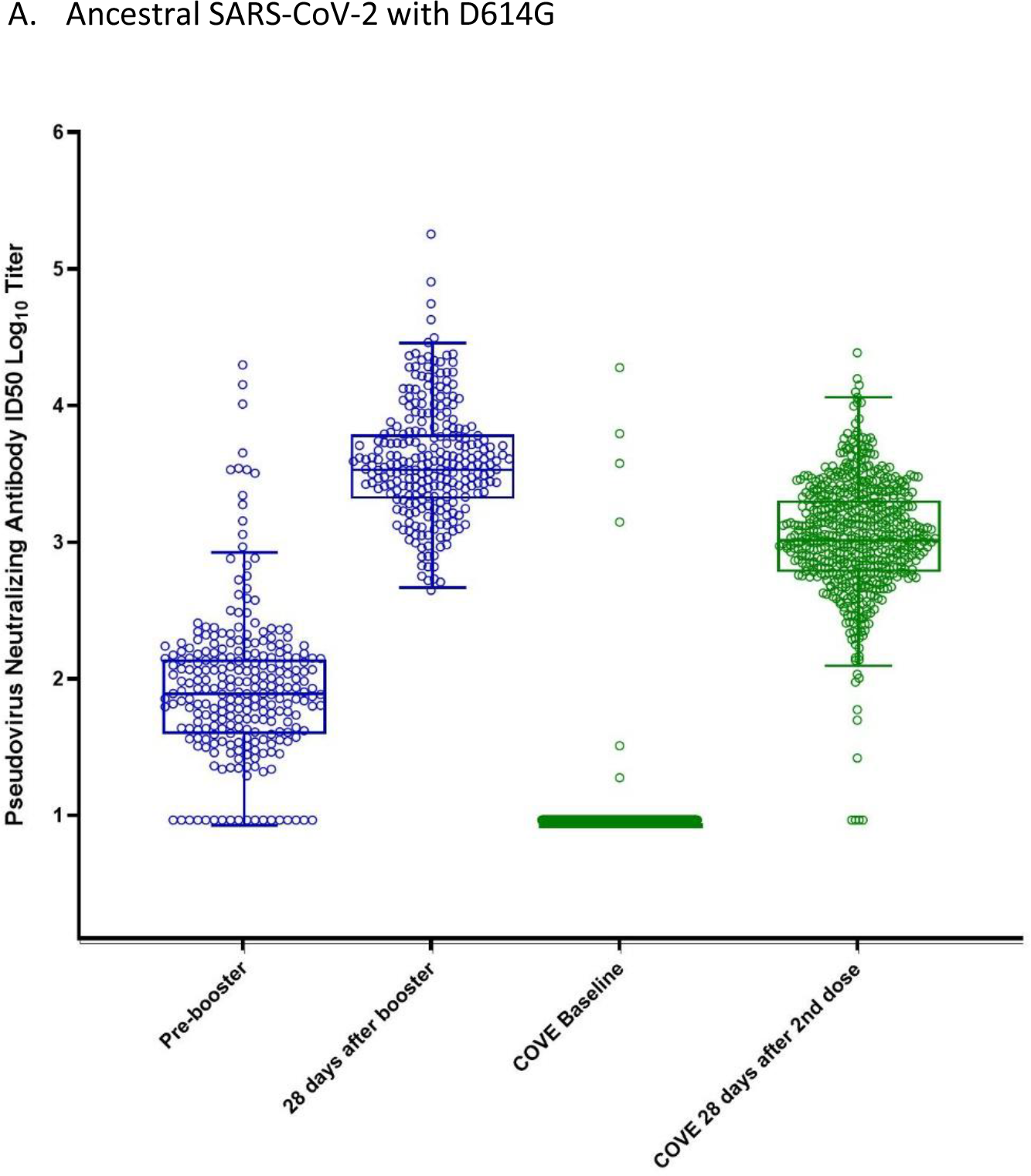
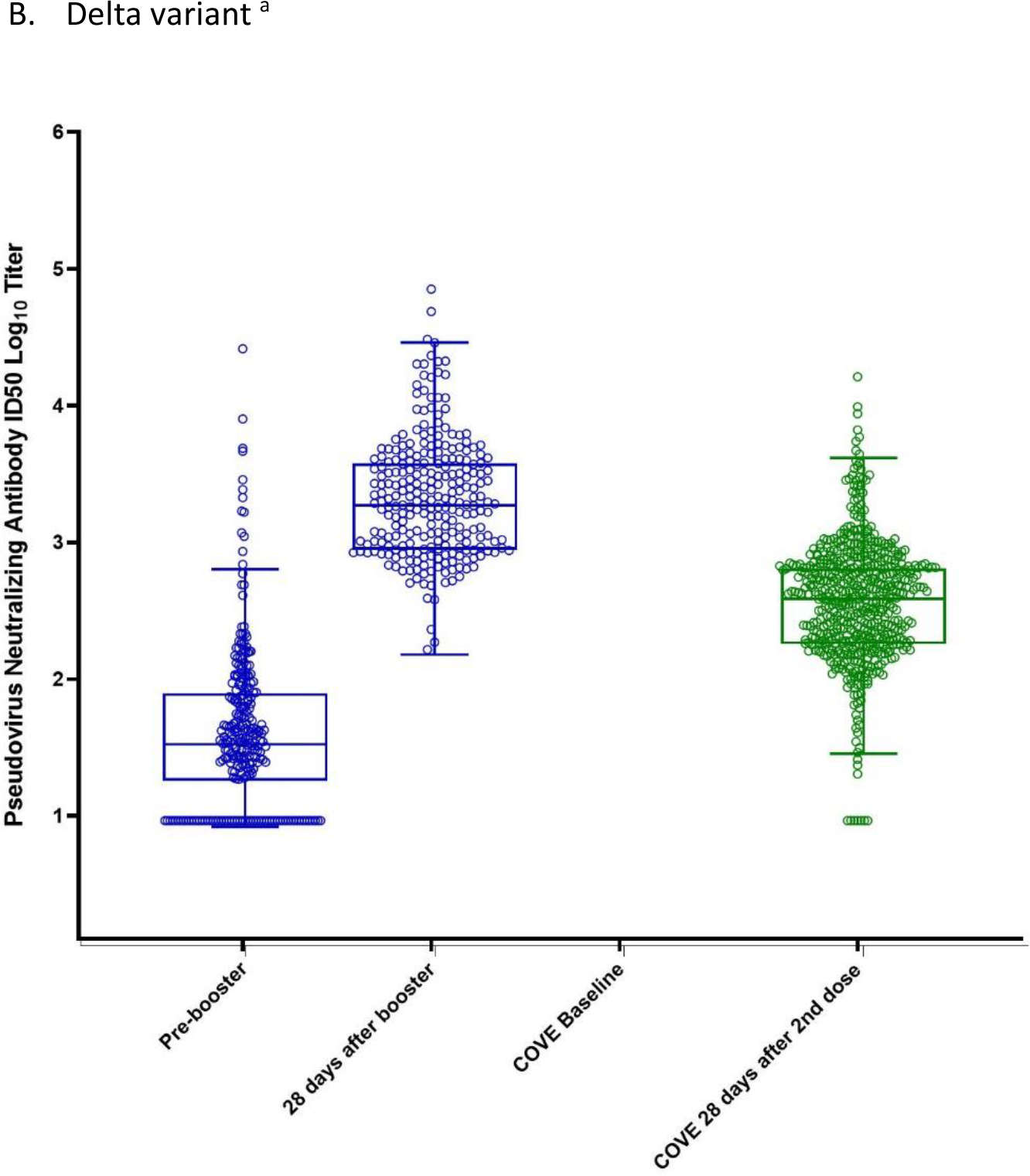
Neutralizing antibody titers (pseudovirus assay; ID50) before and 28 days after the 100 µg booster of mRNA-1273 or the second dose of mRNA-1273 in the phase 3 COVE trial. The neutralizing antibody titers in the pseudovirus assay against the D614G virus (panel A) and against the Delta variant (panel B) are shown for serum samples collected pre-booster and 28 days after the 100 µg mRNA-1273 booster (blue) and from the historical control group (COVE, Phase 3) at baseline and 28 days after the second dose of mRNA-1273 (green). The dots show the results from individual serum samples. The horizontal lines in the middle of the boxes show the median titers. The boxes extend from the 25^th^ percentile to the 75^th^ percentile. The whiskers were determined using the Tukey method. The tops of the whiskers show the 75th percentile plus 1.5 times the IQR (the difference between the 25th and 75th percentiles). The bottoms of the whiskers show the 25th percentile minus 1.5 times the IQR. ^a^ The Delta-specific neutralizing antibody titers were not measured at COVE baseline. The Delta-specific neutralizing antibody titers at the pre-vaccination baseline (COVE baseline) for the historical control group were imputed based on the pre-dose 1 SARS-CoV-2 status; when SARS-CoV-2 status was negative, antibody titers were imputed as <LLOQ at pre-dose 1 of primary series.

Pseudovirus neutralizing antibody titers against the Delta variant (B.1.617.2) also increased after the administration of a booster dose of 100 µg mRNA-1273 to participants who previously received two doses of mRNA-1273. (Table 2; Figure 3B; Supplementary Figure 1; Supplementary Tables 2-4). The Delta-specific GMTs (95% CIs) were 9.3 (NE, NE) pre-vaccination and 2164.4 (1915.0,2446.3) at 28 days after the booster dose. The GMFR (95% CIs) for the Delta-specific GMTs at 28 days after the booster was 233.3 (206.4, 263.7) compared to pre-vaccination baseline titers and 48.7 (41.4, 57.3) compared to pre-booster baseline titers. In comparison, the GMT (95% CI) in the historical control group was 383.1 (352.4,416.4) at 28 days after the second dose. The GMR for the GLSM of the titers at 28 days after the booster dose compared to 28 days after the second 100 µg dose was 5.7 (4.9, 6.6). The Delta-specific GMTs 28 days after the 100 µg mRNA-1273 booster was also numerically higher compared to the GMTs after the 50 µg mRNA-1273 booster [827.8 (738.5, 927.9)].^15^

The seroresponse rates (95% CI) for the ancestral virus with D614G were 100% (98.6, 100) at 28 days after the booster dose and 98.1% (96.7,99.1) at 28 days after the second dose (Table 2) and the percentage difference was 1.9 % (0.4, 3.3) with the lower bound of the 95% CI being above –10%, which met the prespecified non-inferiority criterion. The seroresponse rates for the Delta variant were 100% (98.6, 100) at 28 days after the booster dose and 96.2% (94.3, 97.6) at 28 days after the second dose with a percentage difference of 3.8% (2.3, 5.7). The seroresponse rates based on the pre-booster definition yielded consistent results (Supplementary Tables 2 and 4).

## DISCUSSION

The results of this study indicate that a 100 µg booster dose of mRNA-1273 has an acceptable safety profile and a robust antibody response against SARS-CoV-2 and variants of concern. These results are consistent with other studies that have evaluated the 100 µg dose administered as a booster in recipients of the mRNA-1273 primary series and in individuals who underwent primary immunization with vaccines from other manufacturers^16, 17^.

The incidence of local or systemic adverse reactions was overall higher after the 100 µg booster injection than has been observed in previous studies after the second dose of the primary series and the authorized 50 µg mRNA-1273 booster dose.^5, 7^ These comparisons must be made with caution, as the two booster dose levels of mRNA-1273 (100 µg and 50 µg) were not evaluated head-to-head in the same study, study participants were aware they were receiving a higher booster dose and the interval between the primary series and the 100 µg booster dose (∼11 months) was different compared to the interval for the 50 µg booster dose (∼7.2 months)^7^.

Neutralizing antibody titers (pseudovirus) against the ancestral (Wuhan-HU-1) SARS-CoV-2 with the D614G mutation approximately a month after the 100 µg mRNA-1273 booster injection were observed to be >3-fold significantly higher than the titers a month after the second dose of mRNA-1273 during the primary series (COVE study) and the primary non-inferiority immunogenicity objective of the present study was met. This higher level of antibody titers after the booster injection compared to after the second injection is indicative of a robust immune memory response likely due to stimulation of memory B cells.^18–20^ A robust neutralizing antibody response against the SARS-CoV-2 Delta variant was also observed with a 5.7-fold rise in titer levels, 28 days after the booster dose compared to the titers after the second dose in the primary series.

The neutralizing antibody titers against the ancestral SARS-CoV-2 and the Delta variant following the 100 µg booster dose were higher than the titers after the authorized 50 µg booster dose which was evaluated in a separate phase 2 study^7^. The two different dose levels (50 and 100 µg) were not evaluated head-to-head in the same study as a pre-specified objective that would enable direct comparison of the antibody response and were administered in different intervals from the primary series of mRNA-1273 which could also influence the neutralizing antibody titer levels post-boost. The authorized 50 µg booster of mRNA-1273 elicited a robust antibody response beyond the range of antibody titers predictive of protection against COVID-19 based on correlates-of-risk-and-protection analyses against Wuhan-Hu-1 and it has demonstrated improved vaccine effectiveness based on real-world data. ^6, 21, 22^

The highly transmissible and divergent SARS-CoV-2 Omicron variant has recently caused a concerning wave of infections globally. Sera from participants of the present study who received the 100 µg booster dose, as well as sera from recipients of the mRNA-1273 primary series and of the 50 µg booster dose in other studies, were assessed in an Omicron-specific lentivirus neutralization assay and the results were presented elsewhere^12^. Specifically, compared to post-dose 2 and pre-booster levels, Omicron-specific neutralizing antibody titers increased after the 100 µg mRNA-1273 booster dose, albeit at somewhat reduced levels in comparison to the ancestral SARS-CoV-2. In addition, Omicron-specific titers were numerically higher after the 100 µg booster dose compared to the 50 µg booster dose.

There are several limitations of this study. The evaluation of a 100 µg booster was open-label, and the study was not designed to evaluate booster doses at varying intervals or compare different booster dose levels head-to-head. Additionally, neutralization results from different groups were not generated in the lab at the same time, the historical control group is not the same group of participants enrolled in the present study, and the study was not designed to evaluate vaccine effectiveness. However, neutralizing antibody responses have been correlated to reduction of risk for breakthrough COVID-19.^12, 21, 22^

The strengths of the study include that over 300 participants received a booster dose, allowing for a robust evaluation of possible adverse reactions, and the antibody response against SARS-CoV-2 variants was evaluated. The same laboratory was used to perform all neutralizing antibody assessments in a formally validated assay.

The results from this study provide evidence that a 100 µg mRNA-1273 booster dose administered at least 6 months after the primary series induces a robust immune response against the ancestral SARS-CoV-2 and against variants of concern. The results also suggest that the 100 µg booster dose enhances the breadth of the neutralizing antibody response and elicits even numerically higher neutralizing antibody titers against antigenically divergent variants compared to a 50 µg booster dose. Although observed to have a clinically-acceptable safety profile, both the frequency and severity of reported solicited adverse events with the 100 µg were higher than after the 50 µg booster dose.

The clinical implications of the higher antibody responses observed with the 100 µg booster dose are not known, but the mRNA-1273 100 µg booster dose might be considered in scenarios where eliciting an antibody response might be particularly challenging. Such scenarios include moderately or severely immunocompromised hosts as well emergent variants with antibody escape mutations and evidence of substantially reduced or minimal neutralization with the 50 µg booster dose. Continued monitoring of real-world data of booster doses is needed to assess their long-term effectiveness.

## Funding

The phase 2/3 study (Phase 2/3 Study to Evaluate the Immunogenicity and Safety of mRNA Vaccine Boosters for SARS CoV-2 Variants; NCT04927065) was funded by Moderna, Inc. The Duke laboratory received funding for sample analysis from Moderna, Inc. The Coronavirus Efficacy (COVE) trial (NCT04470427) was supported by the Office of the Assistant Secretary for Preparedness and Response, Biomedical Advanced Research and Development Authority (contract 75A50120C00034) and by the National Institute of Allergy and Infectious Diseases (NIAID). The NIAID provides grant funding to the HIV Vaccine Trials Network (HVTN) Leadership and Operations Center (UM1 AI 68614HVTN), the Statistics and Data Management Center (UM1 AI 68635), the HVTN Laboratory Center (UM1 AI 68618), the HIV Prevention Trials Network Leadership and Operations Center (UM1 AI 68619), the AIDS Clinical Trials Group Leadership and Operations Center (UM1 AI 68636), and the Infectious Diseases Clinical Research Consortium leadership group 5 (UM1 AI148684-03). Parts A and B of the phase 2 trial (Dose-Confirmation Study to Evaluate the Safety, Reactogenicity, and Immunogenicity of mRNA-1273 COVID-19 Vaccine in Adults Aged 18 Years and Older; NCT04405076) were supported in part with Federal funds from the Office of the Assistant Secretary for Preparedness and Response, Biomedical Advanced Research and Development Authority, under Contract No. 75A50120C00034, and Moderna, Inc.

## Data Availability

As the trial is ongoing, access to patient-level data presented in this article (antibody assays, safety and reactogenicity) and supporting clinical documents with external researchers who provide methodologically sound scientific proposals will be available upon reasonable request and subject to review once the trial is complete. Such requests can be made to Moderna Inc., 200 Technology Square, Cambridge, MA 02139. A materials transfer and/or data access agreement with the sponsor will be required for accessing shared data. All other relevant data are presented in the paper. The protocol is available as online supplementary material to this article. ClinicalTrials.gov: NCT04927065.

## Acknowledgements

We thank the participants in this trial, the study investigators, and the members of the Immune Assay Team at Duke University Medical Center. We thank Joanne E. Tomassini, consultant to Moderna, Inc., for her contributions to the manuscript and Barbara Kuter, consultant to Moderna, Inc., for her critical review of the manuscript.

## SUPPLEMENTARY APPENDIX

### LIST OF INVESTIGATORS

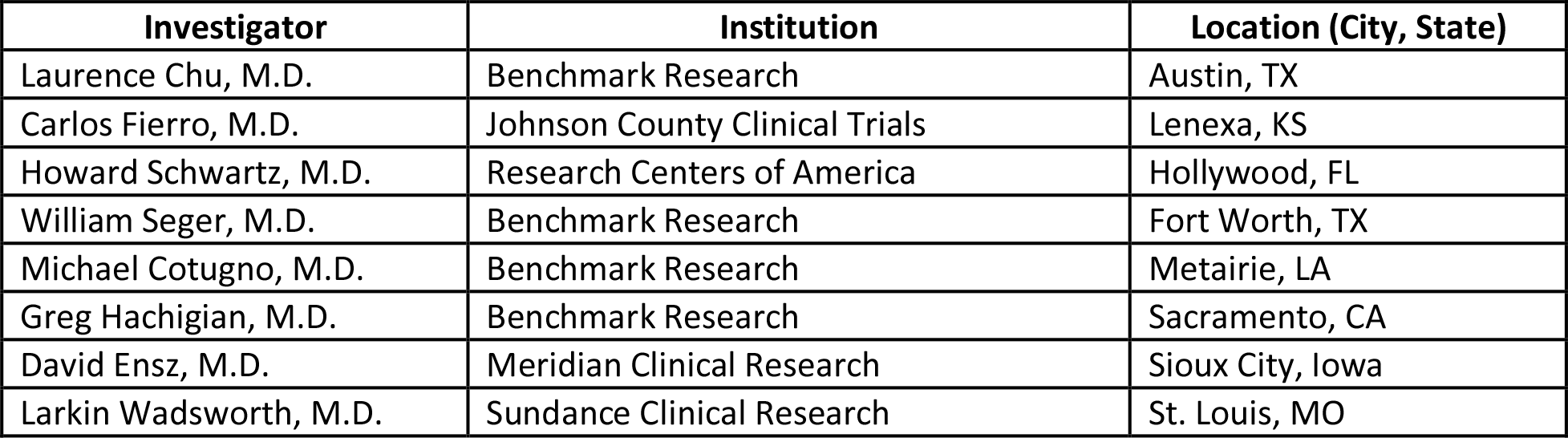

### SUPPLEMENTARY METHODS

#### Immunogenicity assays

##### SARS-CoV-2 Spike-Pseudotyped Virus Neutralization Assay

The SARS-CoV-2 spike-pseudotyped virus neutralization assay was used to analyze all of the samples from this study as well as the historical control group (Phase 3 COVE trial). This is a validated assay that quantifies SARS-CoV-2 neutralizing antibodies by using lentivirus particles that express SARS-CoV-2 full-length spike proteins (Wuhan-Hu-1 isolate including the amino acid change of D614G in the spike protein; the Delta variant [B.1.617.2; AY.3; Wuhan-Hu-1 isolate containing spike mutations T19R, G142D, Δ156- 157, R158G, L452R, T478K, D614G, P681R, D950N] on their surface and contain a firefly luciferase reporter gene for quantitative measurements of infection by relative luminescence units (RLU). The Omicron variant (B.1.1.529) assay and results are described elsewhere^12^.

##### SARS-CoV-2 Meso-Scale Discovery (MSD) assay

The validated Meso-Scale Discovery (MSD) assay uses an indirect, quantitative, electrochemiluminescence method to detect SARS-CoV-2 binding IgG antibodies to the SARS-CoV-2 full-length spike (Wuhan-Hu-1 ancestral isolate including D614G; Beta [B.1.351] with the following amino acid changes in the spike protein [L18F, D80A, D215G, Δ242-244, R246I, K417N, E484K, N501Y, D614G, and A701V]) in human serum. The assay is based on the MSD technology which employs capture molecule MULTI-SPOT® microtiter plates fitted with a series of electrodes.

#### Statistical analysis

Study analysis populations included the Safety Set, comprising all enrolled participants who received the 100 µg mRNA-1273 booster injection; the Immunogenicity Set, consisting all participants who received the booster dose and had neutralizing antibody data at both pre-booster baseline and Day 29 visit, with no major protocol deviations that could impact data integrity. The Safety Set was used for the safety analyses and the Immunogenicity Set and the analysis of immunogenicity data.

The primary immunogenicity objective for 100 µg mRNA-1273 booster dose was considered to be met if the pre-specified non-inferiority criteria were demonstrated, at a 2-sided alpha of 0.05, based on both GMT and SRR at 28 days after the booster dose compared to GMTs 28 days after the second dose in the historical control arm. For the primary immunogenicity objective, there were two pre-specified hypotheses to be tested: (1) Non-inferiority based on the GMR (GMT post-boost/GMT post-primary series) with a noninferiority margin of 1.5; if the lower bound of the 95% CI of GMR ≥ 0.67 (1/1.5), then non-inferiority was considered met; and (2) noninferiority based on the difference in seroresponse rate (SRR) with a noninferiority margin of 10%; if the lower bound of the 95% CI of SRR difference >-10%, then SRR non-inferiority was considered met. Seroresponse was defined as: 4-fold rise for participants with baseline ≥ lower limit of quantification (LLOQ) of the neutralizing antibody assay or ≥ 4 times the LLOQ of the assay for those with baseline <LLOQ. Seroresponse was derived based on two types of baselines: pre-vaccination baseline on Day 1 of the primary series (before receiving the first dose), and pre-booster baseline. The seroresponse rate based on the pre-vaccination baseline titers is the primary definition. For participants who received the 100 µg booster of mRNA-1273 in this study and did not have pre-vaccination antibody titers, seroresponse was defined as ≥ 4 times LLOQ for subjects with negative SARS-CoV-2 status at their pre-dose 1 of primary series, and these participant’s antibody titers were deemed <LLOQ at pre-dose 1 of primary series.

Analysis of covariance (ANCOVA) model was used to assess immune response of 100 µg mRNA-1273 booster dose compared to the immune response after the second dose of 100 µg primary series. The model included log-transformed antibody titers at 28 days post-boost and 28 days post-second dose as the dependent variables, treatment groups (100 µg mRNA-1273 booster dose, 100 µg primary series) as explanatory variable and adjusting for age groups (< 65 years; ≥ 65 years). The geometric least squares mean (GLSM) and corresponding 2-sided 95% CI for the antibody titers for each treatment group were calculated. The GLSM, and the corresponding 95% CI results in log-transformed scale estimated from the model were back-transformed to obtain these estimates in the original scale. GMR, estimated by the ratio of GLSM and the corresponding 2-sided 95% CI were used to assess the treatment difference.

To assess noninferiority of immune response based on SRR, the number and percentage (rate) of participants achieving seroresponse at 28 days after the booster or after the second dose in the historical control group were summarized with 95% CI calculated using the Clopper-Pearson method for each group. The difference of SRRs between 100 µg mRNA-1273 at 28 days after the booster dose and 100 µg mRNA-1273 primary 28 days after the second dose were calculated with 95% CI using Miettinen-Nurminen (score) method.

All analyses were conducted using SAS Version 9.4 or higher.

### IMMUNOGENICITY RESULTS

Administration of a booster dose of 100 µg mRNA-1273 to participants who previously received the primary series of 100 µg mRNA-1273 increased neutralizing antibodies (nAbs) (pseudovirus assay) based on the pre-booster baseline to the ancestral SARS-CoV-2 spike protein containing the D614G amino acid substitution and the Delta variant (B.1.617.2) spike protein (Supplementary Table 2). The GMTs (95% CIs) pre-booster for nAbs to the ancestral virus spike protein were 90.0 (76.9, 104.7) and increased to 4039.5 (3592.7, 4541.8) at 28 days after the booster dose. The GMFR was 45.0 (38.9, 52.1) compared to the pre-booster. In comparison, the GMT (95% CI) for the historical control group was 1132.0 (1046.7, 1224.2) at 28 days after the second dose. The seroresponse rates (95% CI) for the ancestral spike protein were 96.5% (93.5, 98.4) for the booster dose and 98.1% (96.7, 99.1) for the historical control group. Testing for nAbs was also performed against the Delta variant (B.1.617.2) spike protein. The Delta-specific GMTs (95% CI) for nAbs was 2164.4 (1915.0, 2446.3) at 28 days after the booster and 383.1 (352.4, 416.4) in the historical control group at 28 days after the second dose.

Administration of a booster dose of 100 µg mRNA-1273 to participants who previously received the primary series of 100 µg mRNA-1273 also increased binding IgG antibodies (MSD assay) based on the pre-vaccination baseline to the ancestral SARS-CoV-2 spike protein containing the D614G amino acid substitution and the Beta variant (B.1.351) spike protein (Supplementary Table 3). The GMTs (95% CIs) pre-vaccination for binding antibodies to the ancestral virus spike protein were 12 (not estimated [NE], NE) pre-vaccination and increased to 712,284 (577,541-878,464) at 28 days after the booster dose. The GMFR (95% CIs) for the GMTs at 28 days after the booster was 59,469 (48,153-73,444). In comparison, the GMT (95% CI) for the historical control group was 249,485 (229,020-271,779) at 28 days after the second dose.

The seroresponse rates (95% CI) for the ancestral spike protein were 98% (96-99) for the booster dose and 99% (98,100) for the historical control group. Testing for binding antibodies was also performed against the Beta variant (B.1.351) spike protein. The Beta-specific GMTs (95% CI) for binding antibodies were 306,816 (247,479-380,379) at 28 days after the booster and 98,520 (90,730-106,980) in the historical control group at 28 days after the second dose. Administration of a booster dose of 100 µg mRNA-1273 to participants who previously received the primary series of 100 µg mRNA-1273 also increased binding IgG antibodies (MSD assay) based on the pre-booster baseline to the ancestral SARS-CoV-2 spike protein containing the D614G amino acid substitution and the Beta variant (B.1.351) spike protein (Supplementary Table 4). The GMTs (95% CIs) pre-booster for binding antibodies to the ancestral virus spike protein were 23,972 (21,116-27,215) and increased to 712,284 (577,541-878,464) at 28 days after the booster dose. The GMFRs (95% CIs) at 28 days after the booster was 29.7 (23.6- 37.4) based on pre-booster baseline. In comparison, the GMT (95% CI) for the historical control group was 249,485 (229,020-271,779) at 28 days after the second dose. The seroresponse rates (95% CI) for the ancestral spike protein were 95% (91-97) for the booster dose and 99% (98,100) for the historical control group.

Conflicts of Interest

S.C ., B.N., J.F., Y.C., H.Z., D.K.E., R.P., B.L., J.M.M and R.D. are employees of Moderna, Inc., and might hold stock/stock options in the company. D.M. has received funding from Moderna, Inc., for assaying clinical samples. F.J.D. is a consultant to Moderna, Inc.

---

**Supplementary Figure 1:**
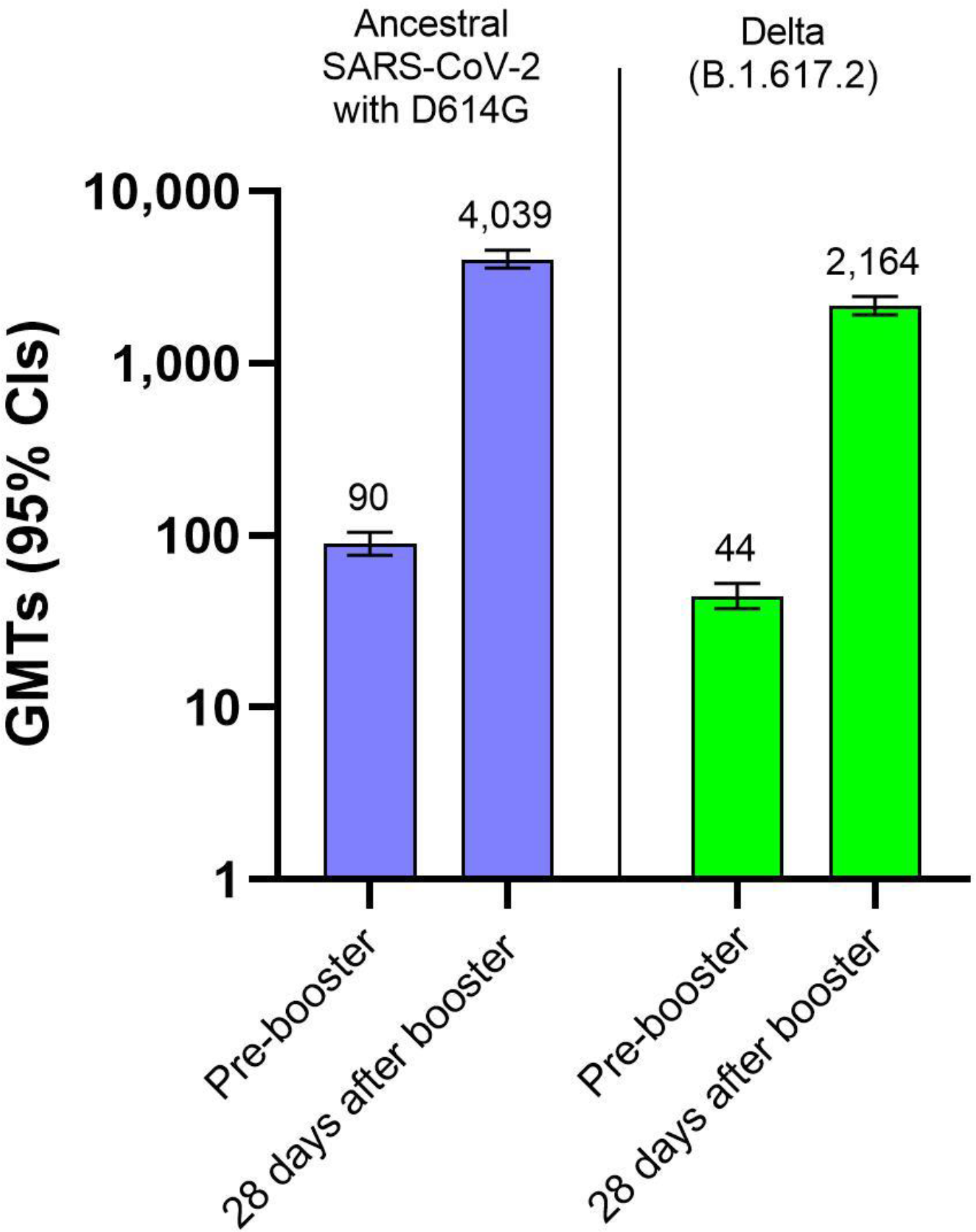
Summary of pseudovirus neutralizing antibodies to SARS-CoV-2 at 28 days after a booster dose of 100 µg of mRNA-1273. GMT=geometric mean titer from participants with non-missing data at the timepoint (pre-booster and 28 days after the booster; CI=confidence interval (95% CI was calculated based on the t-distribution of the log-transformed values or the difference in the log-transformed values for GM value and GM fold-rise, respectively, and then back transformed to the original scale for presentation.)

**SUPPLEMENTARY TABLE 1:** Solicited local and systemic adverse reactions within 7 days after the booster injection of 100 or 50 µg of mRNA-1273 or the second dose of 100 µg of mRNA- 1273. AR=adverse reaction. Any=Grade 1 or higher. N1=Number of exposed participants with any information about the adverse event. Percentages based on N1.

| Adverse Reaction, N (%) | mNA-1273, 100 µg booster, N=303 | mRNA-1273, 50 µg booster in phase 2 mRNA-P201 study N=167 | mRNA-1273 100 µg primary series in Phase 3 COVE study, N=14691 |
| --- | --- | --- | --- |
| Solicited AR, N1 | 303 | 167 | 14691 |
| Any Solicited AR | 290 (95.7) | 151 (90.4) | 13556 (92.3) |
| Grade 1 | 95 (31.4) | 73 (43.7) | 4847 (33.0) |
| Grade 2 | 144 (47.5) | 59 (35.3) | 5800 (39.5) |
| Grade 3 | 51 (16.8) | 18 (10.8) | 2895 (19.7) |
| Grade 4 | 0 | 0 | 14 (<0.1) |
| Any Solicited Local AR, N1 | 302 | 167 | 14688 |
| Any Solicited Local AR | 280 (92.7) | 143 (85.6) | 13029 (88.7) |
| Grade 1 | 183 (60.6) | 108 (64.7) | 8789 (59.8) |
| Grade 2 | 77 (25.5) | 27 (16.2) | 3217 (21.9) |
| Grade 3 | 20 (6.6) | 8 (4.8) | 1023 (7.0) |
| Local AR, Pain, N1 | 302 | 167 | 14688 |
| Pain | 280 (92.7) | 140 (83.8) | 12964 (88.3) |
| Grade 1 | 201 (66.6) | 111 (66.5) | 9508 (64.7) |
| Grade 2 | 68 (22.5) | 23 (13.8) | 2850 (19.4) |
| Grade 3 | 11 (3.6) | 6 (3.6) | 606 (4.1) |
| Erythema, N1 | 302 | 167 | 14687 |
| Erythema | 32 (10.6) | 8 (4.8) | 1274 (8.7) |
| Grade 1 | 14 (4.6) | 5 (3.0) | 456 (3.1) |
| Grade 2 | 14 (4.6) | 2 (1.2) | 531 (3.6) |
| Grade 3 | 4 (1.3) | 1 (0.6) | 287 (2.0) |
| Swelling, N1 | 302 | 167 | 14687 |
| Swelling | 43 (14.2) | 9 (5.4) | 1807 (12.3) |
| Grade 1 | 23 (7.6) | 4 (2.4) | 900 (6.1) |
| Grade 2 | 16 (5.3) | 4 (2.4) | 652 (4.4) |
| Grade 3 | 4 (1.3) | 1 (0.6) | 255 (1.7) |
| Axillary Swelling or<br>Tenderness, N1 | 302 | 167 | 14687 |
| Axillary Swelling | 92 (30.5) | 34 (20.4) | 2092 (14.2) |
| Grade 1 | 79 (26.2) | 30 (18.0) | 1735 (11.8) |
| Grade 2 | 9 (3.0) | 3 (1.8) | 289 (2.0) |
| Grade 3 | 4 (1.3) | 1 (0.6) | 68 (0.5) |
| Any Systemic AR, N1 | 303 | 167 | 14690 |
| Any Systemic AR | 260 (85.8) | 126 (75.4) | 11678 (79.5) |
| Grade 1 | 82 (27.1) | 60 (35.9) | 3717 (25.3) |
| Grade 2 | 140 (46.2) | 53 (31.7) | 5611 (38.2) |
| Grade 3 | 38 (12.5) | 12 (7.2) | 2336 (15.9) |
| Grade 4 | 0 | 0 | 14 (<0.1) |
| Fever, N1 | 303 | 166 | 14682 |
| Fever | 43 (14.2) | 11 (6.6) | 2276 (15.5) |
| Grade 1 | 24 (7.9) | 6 (3.6) | 1363 (9.3) |
| Grade 2 | 16 (5.3) | 3 (1.8) | 697 (4.7) |
| Grade 3 | 3 (1.0) | 2 (1.2) | 203 (1.4) |
| Grade 4 | 0 | 0 | 13 (<0.1) |
| Headache, N1 | 302 | 167 | 14687 |
| Headache | 187 (61.9) | 92 (55.1) | 8637 (58.8) |
| Grade 1 | 105 (34.8) | 61 (36.5) | 4815 (32.8) |
| Grade 2 | 76 (25.2) | 29 (17.4) | 3156 (21.5) |
| Grade 3 | 6 (2.0) | 2 (1.2) | 666 (4.5) |
| Grade 4 | 0 | 0 | 0 |
| Fatigue, N1 | 302 | 167 | 14687 |
| Fatigue | 220 (72.8) | 98 (58.7) | 9607 (65.4) |
| Grade 1 | 81 (26.8) | 47 (28.1) | 3431 (23.4) |
| Grade 2 | 116 (38.4) | 44 (26.3) | 4743 (32.3) |
| Grade 3 | 23 (7.6) | 7 (4.2) | 1433 (9.8) |
| Grade 4 | 0 | 0 | 0 |
| Myalgia, N1 | 302 | 167 | 14687 |
| Myalgia | 204 (67.5) | 82 (49.1) | 8529 (58.1) |
| Grade 1 | 80 (26.5) | 47 (28.1) | 3242 (22.1) |
| Grade 2 | 104 (34.4) | 30 (18.0) | 3966 (27.0) |
| Grade 3 | 20 (6.6) | 5 (3.0) | 1321 (9.0) |
| Grade 4 | 0 | 0 | 0 |
| Arthralgia, N1 | 302 | 167 | 14687 |
| Arthralgia | 147 (48.7) | 69 (41.3) | 6303 (42.9) |
| Grade 1 | 63 (20.9) | 43 (25.7) | 2809 (19.1) |
| Grade 2 | 69 (22.8) | 21 (12.6) | 2719 (18.5) |
| Grade 3 | 15 (5.0) | 5 (3.0) | 775 (5.3) |
| Grade 4 | 0 | 0 | 0 |
| Nausea/ vomiting, N1 | 302 | 167 | 14687 |
| Nausea / vomiting | 59 (19.5) | 19 (11.4) | 2794 (19.0) |
| Grade 1 | 42 (13.9) | 16 (9.6) | 2094 (14.3) |
| Grade 2 | 17 (5.6) | 3 (1.8) | 678 (4.6) |
| Grade 3 | 0 | 0 | 21 (0.1) |
| Grade 4 | 0 | 0 | 1 (<0.1) |
| Chills, N1 | 302 | 167 | 14687 |
| Chills | 133 (44.0) | 59 (35.3) | 6500 (44.3) |
| Grade 1 | 63 (20.9) | 36 (21.6) | 2907 (19.8) |
| Grade 2 | 67 (22.2) | 23 (13.8) | 3402 (23.2) |
| Grade 3 | 3 (1.0) | 0 | 191 (1.3) |
| Grade 4 | 0 | 0 | 0 |

**Supplementary Table 2.** Neutralizing antibody titers (pseudovirus assay; ID50; pre-booster baseline titers) after the 100 µg booster dose of mRNA-1273. GM=geometric mean, CI=confidence interval, GMFR=geometric mean fold rise, GLSM=geometric least squares mean, GMR=geometric mean ratio. ^a^ Number of participants with non-missing data at the corresponding timepoint. ^b^The 95% CI was calculated based on the t-distribution of the log-transformed values or the difference in the log-transformed values for GM value and GM fold-rise, respectively, and then back transformed to the original scale for presentation. ^c^Seroresponse at a participant level was defined as a change from below the LLOQ to equal or above 4 x LLOQ if the baseline was below the LLOQ, or at least a 4-fold rise if the baseline was equal to or above the LLOQ. ^d^The number of participants meeting the criterion at the time point. Percentages are based on N1. ^e^The 95% CI was calculated using the Clopper-Pearson method. ^f^The 95% CI was calculated using the Miettinen-Nurinen (score) confidence limits. For calculation of GMTs and GMFRs, antibody values reported as below the lower limit of quantification (LLOQ) were replaced by 0.5 times the LLOQ. Values that were greater than the upper limit of quantification (ULOQ) were converted to the ULOQ if actual values were not available. Missing results were not imputed.

|  | Ancestral SARS-CoV-2 with D614G |  | Delta (B.1.617.2) |  |
| --- | --- | --- | --- | --- |
|  | Recipients of 100 µg mRNA-1273 Booster Dose<br>N=257 | Historical control group<br>100 µg mRNA-1273<br>N=584 | Recipients of 100 µg mRNA-1273 Booster Dose<br>N=257 | Historical control group<br>100 µg mRNA-1273<br>N=584 |
| Pre-booster/Baseline<br>n<sup>a</sup> | 257 | 584 | 257 |  |
| GM titer | 90.0 | 9.7 | 44.4 |  |
| (95% CI)<sup>b</sup> | (76.9-104.7) | (9.3-10.1) | (37.5-52.6) |  |
| 28 days after booster or 2<sup>nd</sup> dose<br>n<sup>a</sup> | 257 | 584 | 257 | 580 |
| GM titer | 4039.5 | 1132.0 | 2164.4 | 383.1 |
| (95% CI)<sup>b</sup> | (3592.7-4541.8) | (1046.7-1224.2) | (1915.0-2446.3) | (352.4-416.4) |
| GMFR | 45.0 | 116.8 | 48.7 |  |
| (95% CI)<sup>b</sup> | (38.9-52.1) | (107.2-127.3) | (41.4, 57.3) |  |
| Seroresponse rate<sup>±c</sup> |  |  |  |  |
| n/N1 (%)<sup>d</sup> | 248/257 (96.5) | 573/584 (98.1) | 245/257 (95.3) |  |
| (95% CI)<sup>e</sup> | 93.5-98.4 | (96.7-99.1) | (92.0-97.6) |  |
| Difference in seroresponse rate after booster compared to Phase 3 COVE [%; (95% CI)<sup>f</sup>] | -1.6<br>(-4.8, 0.6) |  |  |  |

**Supplementary Table 3.** Binding IgG antibody titers (MSD assay; ID50; Ancestral SARS-CoV-2 with D614G and Beta (B.1.351); pre-vaccination baseline titers) after the 100 µg booster dose of mRNA- 1273. GM=geometric mean, NE=not estimated, CI=confidence interval, GMFR=geometric mean fold rise. ^a^ Number of participants with non-missing data at the timepoint (baseline or post-baseline). ^b^AU/mL. ^c^The 95% CI was calculated based on the t-distribution of the log-transformed values or the difference in the log-transformed values for GM value or GM fold-rise, respectively, then back transformed to the original scale for presentation. ^d^ Seroresponse at a participant level was defined as a change from below the LLOQ to equal or above 4 x LLOQ if the baseline was below the LLOQ, or at least a 4-fold rise if the baseline was equal to or above the LLOQ. For mRNA1273-P205 subjects without pre-Dose 1 antibody titer information and have corresponding Day 29 post-boost assessment, seroresponse is defined as >= 4*LLOQ for subjects with negative SARS-CoV-2 status at their pre-dose 1 of primary series, and these subjects’ antibody titer are imputed as <LLOQ at pre-dose 1 of primary series. For subjects who are without SARS-CoV-2 status information at pre-dose 1 of primary series, their pre-booster SARS-CoV-2 status is used to impute their SARS-CoV-2 status at their pre-dose 1 of primary series. ^e^The number of participants meeting the criterion at the time point. Percentages are based on N1. ^f^The 95% CI was calculated using the Clopper-Pearson method.

|  | Ancestral SARS-CoV-2 with D614G |  | Beta (B.1.351) |  |
| --- | --- | --- | --- | --- |
|  | Recipients of 100 µg mRNA-1273 Booster Dose<br>N=257 | Historical control group, 100 µg mRNA-1273<br>N=584 | Recipients of 100 µg mRNA-1273 Booster Dose<br>N=257 | Historical control group, 100 µg mRNA-1273<br>N=584 |
| Pre-Vaccination |  |  |  |  |
| Baseline n<sup>a</sup> | 255 | 581 | 255 | 582 |
| GM titer<sup>b</sup> | 12 | 44 | 9 | 34 |
| (95% CI)<sup>c</sup> | (NE-NE) | (41-49) | (NE-NE) | (31-37) |
| 28 days after booster or 2<sup>nd</sup> dose<br>n* | 257 | 584 | 257 | 583 |
| GM titer<sup>b</sup> | 712, 284 | 249,485 | 306,816 | 98,520 |
| (95% CI)<sup>c</sup> | (577,541-878,464) | (229,020-271,779) | (247,479-380,379) | (90,730-106,980) |
| GMFR | 59,469 | 5,611 | 34,123 | 2,885 |
| (95% CI)<sup>c</sup> | (48,153-73,444) | (4,953-6,356) | (27,484-42,365) | (2,547-3,267) |
| Seroresponse rate<sup>d</sup> |  |  |  |  |
| n/N1 (%)<sup>e</sup> | 250/255 (98) | 577/581 (99) | 249/255 (98) | 577/581 (99) |
| (95% CI)<sup>f</sup> | 96-99 | (98-100) | (95-99) | (98-100) |

**Supplementary Table 4.** Binding IgG antibody titers (MSD assay; ID50; Ancestral SARS-CoV-2 with D614G and Beta (B.1.351); pre-booster baseline titers) after the 100 µg booster dose of mRNA-1273. GM=geometric mean, CI=confidence interval, GMFR=geometric mean fold rise. ^a^Number of participants with non-missing data at the timepoint (baseline or post-baseline). ^b^AU/mL. ^c^The 95% CI was calculated based on the t-distribution of the log-transformed values or the difference in the log-transformed values for GM value or GM fold-rise, respectively, then back transformed to the original scale for presentation. ^d^Seroresponse at a participant level was defined as a change from below the LLOQ to equal or above 4 x LLOQ if the baseline was below the LLOQ, or at least a 4-fold rise if the baseline was equal to or above the LLOQ. ^e^The number of participants meeting the criterion at the time point. Percentages are based on N1. ^f^The 95% CI was calculated using the Clopper-Pearson method.

|  | Ancestral SARS-CoV-2 with D614G |  | Beta (B.1.351) |  |
| --- | --- | --- | --- | --- |
|  | Recipients of 100 µg mRNA-1273 Booster Dose<br>N=257 | Historical control group, 100 µg mRNA-1273<br>N=584 | Recipients of 100 µg mRNA-1273 Booster Dose<br>N=257 | Historical control group, 100 µg mRNA-1273<br>N=584 |
| Pre-booster/Baseline n<sup>a</sup> | 257 | 581 | 257 | 582 |
| GM titer<sup>b</sup> | 23,972 | 44 | 8,973 | 34 |
| (95% CI)<sup>c</sup> | (21,116-27215) | (41-49) | (7,904-10,185) | (31-37) |
| 28 days 9 after booster or 2<sup>nd</sup> dose<br>n<sup>*</sup> | 257 | 584 | 257 | 583 |
| GM titer<sup>b</sup> | 712,284 | 249,485 | 306,816 | 98,520 |
| (95% CI)<sup>c</sup> | (577,541-878,464) | (229,020-271,779) | (247,479-380,379) | (90,730-106,980) |
| GMFR | 30 | 5,611 | 34 | 2,885 |
| (95% CI)<sup>c</sup> | (24-37) | (4,953-6,356) | (27-43) | (2,547-3,267) |
| Seroresponse rate<sup>d</sup> |  |  |  |  |
| n/N1 (%)<sup>e</sup> | 243/257 (95) | 577/581 (99) | 244/257 (95) | 577/581 (99) |
| (95% CI)<sup>f</sup> | 91-97 | (98-100) | (92-97) | (98-100) |

## Notes

### Competing Interest Statement

S.C., B.N., J.F., Y.C., H.Z., D.K.E., R.P., B.L., J.M.M and R.D. are employees of Moderna and might hold stock/stock options in the company. D.M. has received funding from Moderna for assaying clinical samples. F.J.D. is a consultant to Moderna.

### Clinical Trial

NCT04927065

### Author Declarations

Protocol was approved by Advarra Inc., Columbia, MD.

